# RT-qPCR half reaction optimization for the detection of SARS-CoV-2

**DOI:** 10.1101/2021.05.19.21257470

**Authors:** Priscila Lamb Wink, Fabiana Volpato, Daiana de Lima-Morales, Rodrigo Minuto Paiva, Julia Biz Willig, Hugo Bock, Fernanda de Paris, Afonso Luís Barth

**Author notes:** Corresponding author: Afonso Luís Barth, LABRESIS- Laboratório de Pesquisa em Resistência Bacteriana, Hospital de Clínicas de Porto Alegre, Rua Ramiro Barcelos 2350, Porto Alegre, RS, Brazil, 90.035-903.

## Abstract

**BACKGROUND:** The main laboratory test for the diagnosis of COVID-19 is the reverse transcription real-time polymerase chain reaction (RT-qPCR). However, the RT-qPCR is an expensive method due to the number of tests required.

**OBJECTIVES:** To evaluate an alternative RT-qPCR approach for the detection of SARS-CoV-2 sing half of the total volume currently recommended by the US Centers for Disease Control and Prevention.

**METHODS:** The analytical limit of detection (LoD) and the reaction efficiency using half volumes of RT-qPCR assay were evaluated for both the N1 and N2 regions by using a synthetic control RNA. A panel of 76 SARS-CoV-2-positive and 26 SARS-CoV-2-negative clinical samples were evaluated to establish the clinical sensitivity and specificity.

**FINDINGS:** The RT-qPCR assay efficiency was 105% for both the half and standard reactions considering the N2 target and 84% (standard) and 101% (half) for N1. The RT-qPCR half reaction LoD for N1 and N2 were 20 and 80 copies/µL, respectively. Clinical sensitivity and specificity were 100%. The half reaction presented a decrease of up to 5.5 cycle thresholds when compared with the standard RT-qPCR.

**CONCLUSIONS:** The use of RT-qPCR half reaction proved to be a feasible and economic strategy for detection of SARS-CoV-2 RNA.

**Sponsorship:** This work was supported by FAPERGS (20/2551-0000265-9) and by Fundo de Incentivo a Pesquisa e Eventos do Hospital de Clínicas de Porto Alegre (FIPE/HCPA) (Project no. 2020-0163).

## 1. Introduction

Coronavirus disease 2019 (COVID-19), caused by the novel severe acute respiratory syndrome coronavirus 2 (SARS-CoV-2), emerged in Wuhan, China, at the end of 2019 and rapidly spread worldwide. This pandemic has resulted in more than 10 million cases all over the world with 508,000 confirmed deaths, which was accompanied by unprecedented public health action (1,2). A proven effective therapy is currently not available for SARS-CoV-2, which reinforces the importance of massive testing to segregate persons in order to limit the spread of SARS-CoV-2 (3).

Reverse transcriptase polymerase chain reaction (RT-qPCR) is the main method used for the detection of SARS-CoV-2 (2). Distinct RT-qPCR testing protocols comprising different probes and primers used in a multi-step PCR workflow were swiftly established and made publicly available by the WHO, and by the Center for Disease Control (CDC) (4,5).

Considering the high infectivity of SARS-CoV-2, it is very important to increase the RT-qPCR testing rate to allow fast and accurate identification of infected people in order to prevent dissemination of the virus. However, the capacity of the laboratories which perform the RT-qPCR for SARS-CoV-2 worldwide is usually overloaded and there is a need for new protocols to reduce costs.

This study aimed to optimize the RT-qPCR method for the detection of SARS-CoV-2 using only half of the total volume of reagents currently recommended by the CDC protocol. In addition, we tested a panel of 76 SARS-CoV-2-positive and 26 SARS-CoV-2 negative clinical samples using the standard assay in order to obtain a preliminary evaluation of clinical sensitivity and specificity of the half reaction.

## 2. Material and Methods

### 2.1. Nucleic acid extraction

RNA was extracted from 600 μL of respiratory specimens using the Abbott mSample Preparation System (Promega, Madison, WI, USA) with an Abbott M2000 instrument (Abbott, Chicago, EUA), following the manufacturer’s instructions. Total nucleic acids were eluted in 80 μL of Abbott mElution buffer.

### 2.2. Reverse Transcriptase quantitative Polymerase Chain Reaction

Two genes of the nucleocapsid protein (N), N1 and N2, were amplified using a set of primers and probes as described by the Centers for Disease Control and Prevention (CDC - USA) in an RT-qPCR assay. Primers and probes were purchased from Integrated DNA Technologies (Coralville, IA, USA). We performed both the standard RT-qPCR and RT-qPCR half reaction assays in parallel using the SuperScript™ III One-Step RT-PCR System (Thermo Fisher Scientific, Massachusetts, EUA). A description of the RT-qPCR assay is available at the CDC Laboratory Information website for COVID-19 (6). Briefly, to perform the standard RT-qPCR reaction, 5 μL isolated RNA sample was mixed with 15 μL of one-step RT-qPCR mix containing 10 μL of 2X Master Mix, 0.4 μL of Platinum enzyme, 1.5 μL each of 10 μM combined primer/probe mix, 0.4 μL of 2.5 μM ROX passive reference dye, 2.7 μL of water, and 5 μL of nucleic acid extract. For the RT-qPCR half reaction, half of the volumes of the reagents listed above were used, without adding water to the mixture. Therefore, our proposed reaction included 6 μL of reagents plus 4 μL of RNA, totaling 10 μL in each well, resulting in a reaction with more template RNA in relation to the reaction mixture (4–6 μL compared to 5–15 μL from the standard RT-qPCR reaction).

We used three control samples in each RT-qPCR run: a positive template control, water as a negative control, and an internal control (human ribonuclease P gene, RNAse P). We conducted both of the assays (standard reaction and the half reaction) in 96-well plates using an Applied Biosystems QuantStudio Real-Time PCR 3 Instrument (Thermo Fisher Scientific, Massachusetts, EUA). Cycling conditions consisted of 30 min at 50°C for reverse transcription, 2 min at 95°C for activation of the Platinum enzyme, and 45 cycles of 15 s at 95°C and 35 s at 55°C. We used the threshold automatically established by the equipment.

A cycle threshold (Ct) value lower than 40 for N1 and N2 targets was reported as PCR positive. The result was considered negative if the Ct was undetectable or greater than 40. The RNAse P was used to monitor nucleic acid extraction, specimen quality and presence of reaction inhibitors.

### 2.3. Standard curve to assess efficiency

Assay specificity was determined using high-titer virus stock as well as clinical samples positive for SARS-CoV-2 using the standard RT-qPCR assay in a Quant Studio Real-Time PCR system (Thermo Fischer Scientific, Massachusetts, EUA). The reaction efficiency was validated for N1 and N2 targets using a standard curve with five points with a serial dilution (from 1 × 10^5^ to 10 copies/µL) of a synthetic control RNA (Integrated DNA Technologies, Coralville, IA, USA) in triplicate. The data of the Ct values of the serial dilutions were plotted against the target concentration (number of the virus copies). We determined the slope of the curve by linear regression and defined the required levels for PCR efficiency ([100 × 10^(−1/slope)^ −1]) and linearity (*R*^2^) of each RT-qPCR target to be 90–110% and >0.95, respectively (7).

### 2.4. Limit of detection (LoD) of RT-qPCR with N1 and N2

To determine the analytical limit of detection (LoD) of the RT-qPCR half reaction assay, we tested 1, 5, 10, 20, 50, 80, and 100 copies of SARS-CoV-2 RNA per μL. The LoD of N1 and N2 targets were independently assessed using serially diluted synthetic control RNA. The calibration curve for the genomic copy number versus Ct value was obtained from the Quant Studio RT-qPCR instrument (Thermo Fischer Scientific, Massachusetts, EUA). A series of 20 parallel reactions per concentration step was prepared and tested by RT-qPCR.

Assay reproducibility was tested using replicated dilutions of the RNA transcripts and intra- and inter-assay variability was evaluated for each dilution point on different days.

### 2.5. Evaluation of clinical specimens

A total of 102 clinical samples from different patients attending at “Hospital de Clínicas de Porto Alegre” in Southern Brazil were obtained by oro/nasopharyngeal swabbing. All samples were submitted to analysis using the standard and the half reaction protocols described above.

### 2.6. Ethical approval

The study was approved by the Ethics Committees from Hospital de Clínicas de Porto Alegre (CAAE: 30767420.2.0000.5327).

## 3. Results

### 3.2 Standard curve to assess efficiency

The reaction efficiency using half of the volume was evaluated for both N1 and N2 regions in comparison to the standard RT-qPCR reaction by using a synthetic control RNA. The RT-qPCR efficiencies regarding the half reaction were 101.2% and 105.7% for N1 and N2 targets, respectively. Conversely, for the standard RT-qPCR reaction, the efficiency values were 84.4% (N1) and 104.7% (N2) (Table 1). The *R*^2^ for each target was found to be higher than 0.95 for both reactions (Figure 1).

**Table 1.** Efficiency of the two RT-qPCR approaches with a dilution series of a synthetic control RNA.

| Target |  | Mean of Ct values<sup>c</sup> |  |  |  |  | Slope<sup>d</sup> | Efficiency (%)<sup>e</sup> |
| --- | --- | --- | --- | --- | --- | --- | --- | --- |
|  |  | 10<sup>5</sup> | 10<sup>4</sup> | 10<sup>3</sup> | 10<sup>2</sup> | 10<sup>1</sup> |  |  |
|  |  | copies/μL | copies/μL | copies/μL | copies/μL | copies/μL |  |  |
| N1 | SdRT-qPCR<sup>a</sup> | 25.5 | 28.4 | 32.2 | 35.5 | 41.4 | -3.763 | 84.4 |
|  | hRT-qPCR<sup>b</sup> | 24.5 | 27.2 | 30.1 | 32.8 | 39.6 | -3.294 | 101.2 |
| N2 | SdRT-qPCR | 25.6 | 29.2 | 32.6 | 35.2 | ND<sup>f</sup> | -3.214 | 104.7 |
|  | hRT-qPCR | 23.3 | 27.2 | 30.0 | 33.0 | ND | -3.192 | 105.7 |

<sup>a</sup> SdRT-qPCR = Standard RT-qPCR
<sup>b</sup> hRT-qPCR = RT-qPCR half reaction
<sup>c</sup>Values shown are mean of triplicate tests.
<sup>d</sup>Slope = Y intercept – slope log10.
<sup>e</sup>Efficiency = $[100 \times 10^{(-1/\text{slope})} - 1]$ .
<sup>f</sup>ND, not-detected.

**Figure 1.**
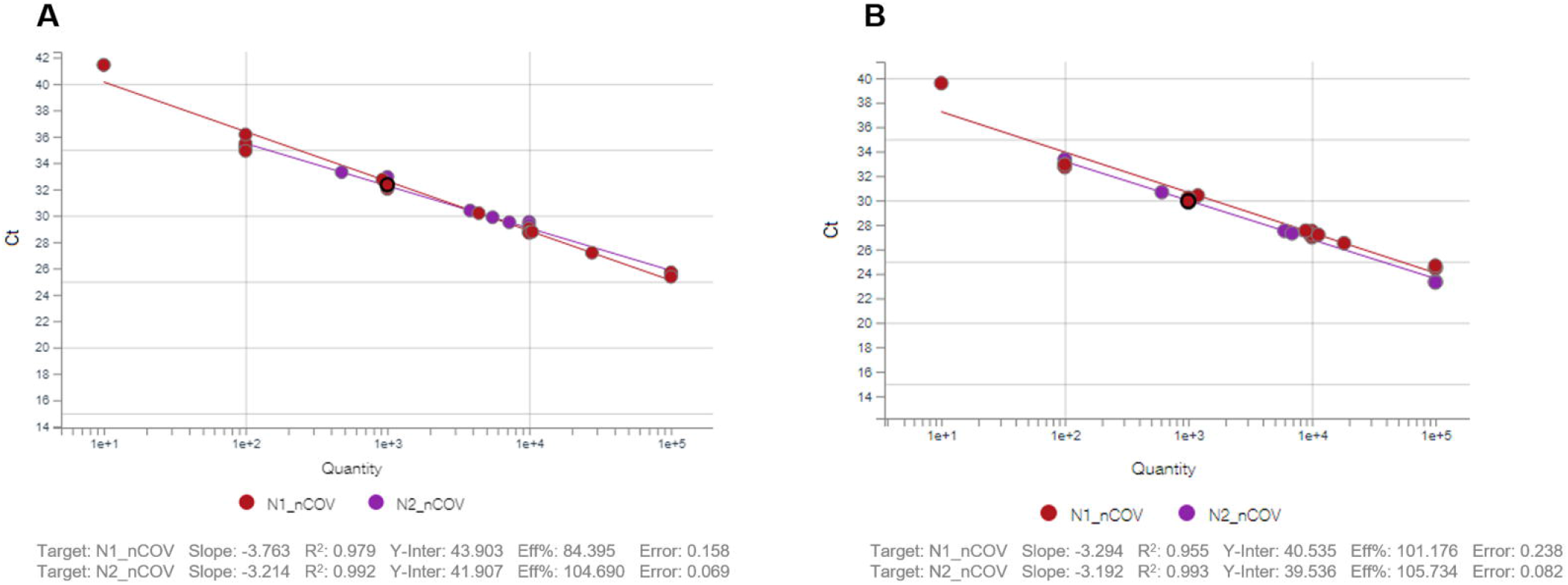
Determination of detection efficiency for N1 and N2 assays. A) RT-qPCR standard curve plot; B) RT-qPCR half reaction standard curve plot. N1_nCOV, N1 target; N2_nCOV, N2 target; R^2^, curve linearity; Eff%, curve efficiency.

Although we planned to generate efficiency curves for both the RT-qPCR half and standard reactions using five dilution points for both targets (N1 and N2), this was only possible for the N1 target. Due to the LoD of the N2 target, we were able to obtain a Ct value for only four dilution points (from 1 × 10^5^ to 1 × 10^2^ copies/µL).

### 3.3 LOD of RT-qPCR with N1 and N2

Serial dilutions of the nucleocapsid RNA transcripts were tested to assess the detection limits and dynamic range of the RT-qPCR assays. The lower LoD was 20 transcript copies per reaction for the N1 target, and 80 copies per reaction for the N2 target.

At these lower copy detection limits for the N1 and N2 targets, assay reproducibility was 95% and 100%, respectively. Eighty-five percent reproducibility was achieved at the dilution that contained 10 transcript copies per reaction with the N1 target and at the 50 transcript copies per reaction dilution with the N2 target.

### 3.2 Evaluation of Clinical Specimens

The RT-qPCR half reaction assay was carried out with 102 clinical samples; all 102 samples were submitted to the standard RT-qPCR described by CDC and 76 samples presented positive results, with Ct values varying from 14.9 to 33.7. The half reaction assay presented positive results for all 76 specimens with positive results with the standard reaction. Likewise, none of the 26 specimens with negative results in the standard reaction presented positive results in the half reaction assay.

The Ct values of the half and standard reactions of RT-qPCR were compared in order to perform a quantitative analysis of the results. The half reaction assay presented a decrease in Ct values for 55 and 73 clinical specimens for the N1 and N2 targets, respectively. The average decrease in Ct values was 0.7 and 2.8 for N1 and N2 targets, respectively, when compared to the standard RT-qPCR (Table 2).

**Table 2.** Comparison of Ct values between the standard and half reactions of RT-qPCR for SARS-CoV-2 positive clinical samples that presented a decrease in the Ct.

| RT-qPCR Ct values | Average of SdRT-qPCR<sup>a</sup> minus hRT-qPCR<sup>b</sup> |  |
| --- | --- | --- |
|  | N1 | N2 |
| ≤20 | 0.5 ± 0.4 (n = 20) | 2.3 ± 0.8 (n = 16) |
| 20-25 | 0.8 ± 0.5 (n = 12) | 2.6 ± 0.8 (n = 21) |
| 25-30 | 0.7 ± 0.5 (n = 16) | 2.7 ± 1.0 (n = 16) |
| >30 | 0.7 ± 0.3 (n = 7) | 3.5 ± 1.3 (n = 20) |
| **All clinical samples** | **0.7 ± 0.4 (n = 55)** | **2.8 ± 1.1 (n = 73)** |

<sup>a</sup>SdRT-qPCR = Standard RT-qPCR
<sup>b</sup>hRT-qPCR = Half reaction RT-qPCR

Twelve isolates with an increase in Ct value for the N1 target (average 0.4) and 2 isolates for the N2 target (0.1) were observed for the half RT-qPCR assay when compared to the standard reaction. Nine isolates did not present a difference in the values of Ct for N1 and only one for the N2 target.

## 4. Discussion

We evaluated RT-qPCR using half of the total reaction volume currently recommended by the US CDC protocol to detect SARS-CoV-2. Whether or not the half reaction assay proved to be a reliable method to detect the RNA of SARS-CoV-2, it would save consumables. The RT-qPCR half reaction efficiency was equivalent to the standard RT-qPCR reaction for the N2 target. Moreover, we observed an important increase in efficiency (from 84.4% to 101.2%) for the N1 target using the RT-qPCR half reaction. The LoD for N1 and N2 was also evaluated for the RT-qPCR half reaction assay and the results demonstrated that N1 is more sensitive than N2 (LoD = 20 and 80 copies/µL, respectively).

These data show that the reduction in the reaction approach has no influence on the quality of the results; unexpectedly, an increase in sensitivity and efficiency was observed. When we compared the Ct values of the SARS-CoV-2 positive clinical samples obtained by the two assays, we observed a decrease in the values of Ct for most samples, either for the N1 (55 cases – 72%) or N2 (73 cases – 96%) targets in the half RT-qPCR assay. These findings indicate that the half reaction assay presents a better yield than the standard reaction. It is possible that the ratio of RNA to reaction mixture could have an influence on the sensitivity of the RT-qPCR. Furthermore, the comparison of Ct values of the two reaction approaches with clinical samples shows the kinetics of sensitivity depending on the viral load using the Ct value. The lower the Ct, the lower the differences between the approaches (Table 2).

Although RT-qPCR assays remain the molecular test of choice for the diagnosis of SARS-CoV-2 infection, considerable efforts have been made to improve the detection of coronavirus and a variety of improved or new approaches have been developed. Since the RT-qPCR methods require time-consuming sample handling and post-PCR analysis, immunoassays have been developed for the rapid detection of SARS-CoV-2 antigens or antibodies (IgM and IgG) against COVID-19. They would theoretically provide the advantage of a fast time to results and the low-cost detection of SARS-CoV-2, but are likely to suffer from poor sensitivity in the early infection period (8).

To enable massive coronavirus testing, the pooling of clinical samples was proposed as a testing strategy that would significantly increase the testing capacity of laboratories (9,10). However, the use of pools would be much more effective to test clinical samples in scenarios of the low prevalence of SARS-CoV-2 as a positive result for the pool would require all samples to be tested individually (9,10). Furthermore, as previously described, considering that the concentrations of RNA had been reduced in the pooled specimens, the Ct values were expected to increase by five Ct values compared with single samples (9). Accordingly, pools with a high number of samples (30 samples) may present false-negative results in the RT-qPCR for SARS-CoV-2, especially when a positive sample has a higher Ct (9).

## 5. Conclusions

The present study optimized the RT-qPCR method for the detection of SARS-CoV-2, with the half reaction presenting a better performance with higher sensitivity and specificity compared to the standard reaction. Importantly, in view of the RT-qPCR reaction being frequently used worldwide at present, it is a huge advantage to adapt this protocol in the routine of molecular diagnostic laboratories, as it is essential for economic purposes to save reagents and increase the number of patients able to be tested for COVID-19 disease worldwide. Moreover, decreasing the total reagent volume and increasing the template concentration inside the reaction assay for a single run would be very useful to increase test sensitivity. In light of the current situation, a one-step PCR protocol achieving both high sensitivity and specificity is beneficial for facing the SARS-CoV-2 pandemic.

## Data Availability

The datasets generated during and/or analyzed during the current study are available from the corresponding author on reasonable request.

## Acknowledgements

We thank the staff of “Laboratório de Diagnóstico de SARS-CoV-2” as well as the staff of the Hospital Infection Committee of our institution (“Hospital de Clínicas de Porto Alegre”) for providing data used in this study.

## Author contributions

A.L.B is the principal investigator of this work and, as such, had full access to all of the data in the study and takes responsibility for the integrity of the data and the accuracy of the data analysis. Author’s contribution: P.L.W. - Study conception and design, acquisition of data, drafting of manuscript, analysis and interpretation of data. F.V. - Analysis and interpretation of data. D.L.M. - Analysis and interpretation of data. R.M.P. - Study conception and design, acquisition of data, analysis and interpretation of data. J.B.W. - Study conception and design, acquisition of data, analysis and interpretation of data. H.B. - Analysis and interpretation of data. F.P. - Analysis and interpretation of data. A.L.B. - Critical revision. All authors critically reviewed.

